# Discrimination, Smoking, and Cardiovascular Disease Risk: A Moderated Mediation Analysis with the Multi-Ethnic Study of Atherosclerosis

**DOI:** 10.1101/2023.09.15.23295645

**Authors:** Stephanie H. Cook, Erica P. Wood, James H. Stein, Robyn L. McClelland

## Abstract

**Background:** Race is a social determinant of cardiovascular (CVD) risk and the American Heart Association has called for increased research to understand how this social determinants of health (SDoH) impacts CVD risk. Carotid intima-media thickness (cIMT) and carotid plaque are reliable indicators of cardiovascular risk. Further, research highlights that disparities exist in these indicators of cardiovascular risk such that racial/ethnic minorities generally exhibit greater characteristics of cardiovascular risk with respect to these indices (e.g., greater cIMT and higher carotid plaque scores) than white individuals due, in part, to exacerbated stress experienced from racial discrimination. At present, the mechanisms driving these racial/ethnic disparities is poorly understood.

**Methods and Results:** Data came from the baseline examination of the Multi-Ethnic Study of Atherosclerosis (MESA). 6,814 participants aged 45-84 free of clinical CVD completed assessments on health behavior, perceived discrimination, CVD-related risk factors, and sociodemographics. Four biological sex-stratified moderated mediation models were used to examine the associations between discrimination, cigarette smoking, and mean cIMT and plaque, modeled separately. We hypothesized that cigarette use would mediate the association between discrimination and carotid artery disease features and that these associations would be moderated by race/ethnicity. While results did not fully support our study hypothesis, racial/ethnic differences were observed across biological sex. Significant indirect effects of discrimination on plaque scores were observed among Hispanic females such that increased discrimination was associated with more cigarette use which, in turn, was associated with higher plaque (*b*=.04, CI=.01, .08). Similar findings were observed among Hispanic males in relation to mean cIMT (*b*=.003, CI=.00, .01) and among white (*b*=.04, CI=.01, .08) and Hispanic males (*b*=.03, CI=.004, .07) and plaque. No other racial/ethnic differences were observed.

**Conclusions:** Results suggest that cardiovascular risk disparities should be examined within frameworks that consider the importance of the intersection of multiple identities (e.g., race and gender). To better address the American Heart Association’s call to examine social determinants of cardiovascular health, researchers must incorporate the complexity of the intersection of various social positions in future work.

## Introduction

The American Heart Association and other prominent health organizations have called for increased research to understand the social determinants health (SDoH) in relation to cardiovascular disease.^1–3^ This work presented within is critically important because it highlights the ways in which the intersection between race and gender drive differences in the association between discrimination, a detrimental SDoH, and known markers of cardiovascular disease – carotid intima-media thickness (cIMT) and carotid plaque. While cardiovascular disease (CVD) remains the leading cause of death among men and women over the age of 65 in the United States,^4,5^ disparities in CVD risk and mortality exist by biological sex and race and ethnicity.^1,6–10^ These disparities may, in part, be explained by social determinants of health such as exposure discrimination. SDoH are defined as societal and environmental conditions that influence health and health behavior of individuals.^11^ Nevertheless, to date, the social determinants of CVD risk across these different gender and racial and ethnic groups are poorly characterized and understood.^2^

Two subclinical CVD measures that are predictive of future cardiovascular events are cIMT and carotid plaque burden,^12–17^ both of which have been shown to differ by characteristics such as race and ethnicity and gender.^18,19^ cIMT and carotid plaque are both measured via B-mode ultrasound and represents a low-burden method through which to measure thickening of the carotid artery as well as buildup of plaque within the artery and are both strong predictors of future cardiovascular events.^12,14,15,17,20,21^ While research that denotes the relevance of discrimination (a social determinant of health) among racial/ethnic minorities in shaping health behaviors and outcomes,^22–26^ the mechanisms linking discrimination and cardiovascular risk factors among racially diverse populations remain poorly understood. Thus, we sought to examine a moderated mediation pathway testing how perceived discrimination relates to health behaviors (i.e., smoking) and, in turn, characteristics of CVD risk including cIMT and carotid plaque burden using the Multi-Ethnic Study of Atherosclerosis. We hypothesized that race and ethnicity would moderate the associations between everyday discrimination and cigarette use in such a way that racial/ethnic minorities will have a stronger positive association between perceived discrimination and the likelihood of cigarette use compared to white males which, in turn, would be associated with higher cIMT and carotid plaque burden (see Figure 1 for our proposed model).

**Figure 1.**
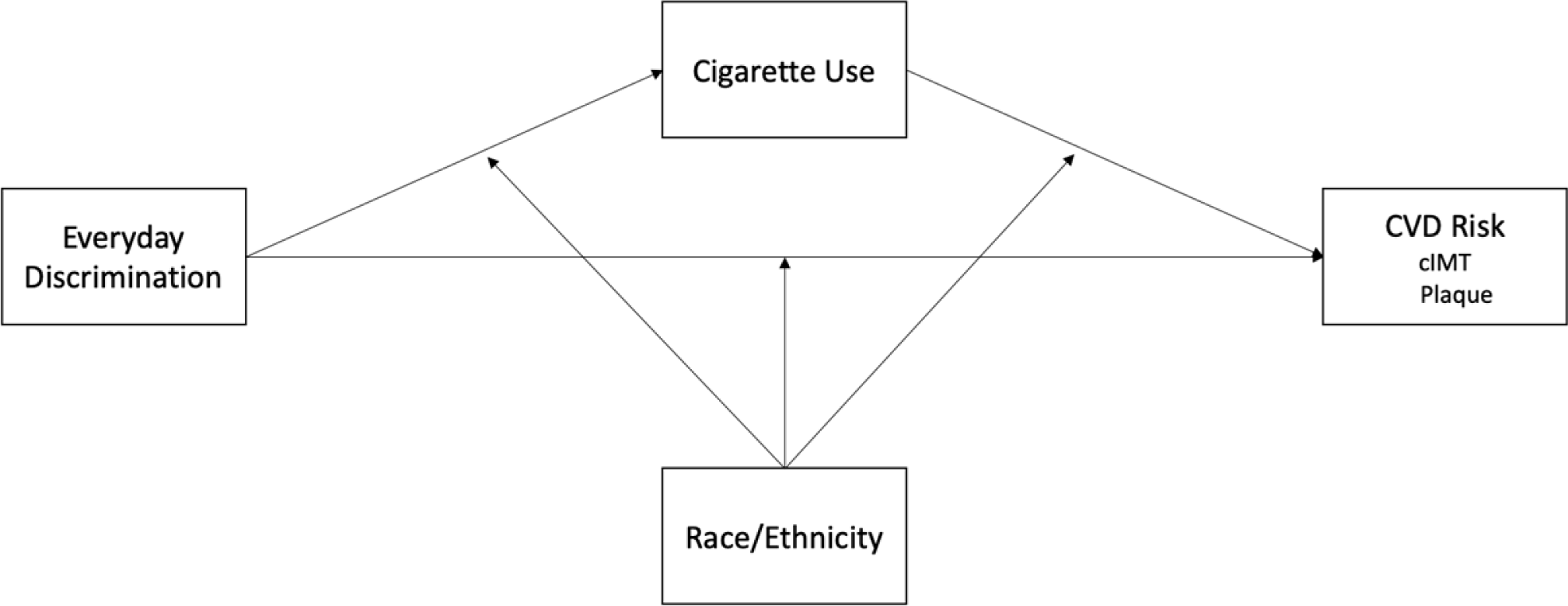
Proposed moderation mediation model

## Methods

### Study Sample

Data came from the Multi-Ethnic Study of Atherosclerosis (MESA), a longitudinal study aimed at examining the characteristics of subclinical CVD as well as factors that contribute to clinical CVD among Americans aged 45-84. Full study details are described elsewhere.^27^ Briefly, this cross-sectional analysis utilized the baseline of MESA data collected between 2000 and 2002 (Wave 1), which included 6,814 participants free of clinical CVD who were recruited from six field centers across the United States (Baltimore, MD; Chicago, IL; St. Paul, MN; Los Angeles, CA; New York, NY; and Forsyth County, NC).

### Study Variables

#### Control Variables

Gender was categorized as sex assigned at birth (female or male), and race was self-identified and represented one of four categories: White American, Chinese-American, African-American, and Hispanic. Several covariates were included due to their known association with CVD risk (i.e., physical activity, blood pressure, glucose, and total cholesterol).^28–31^ Physical activity level was measured at baseline from the MESA Typical Week Physical Activity Survey. This survey, adapted from the Cross-Cultural Activity Participation Study,^32^ examines frequency of and time spent in various physical activities (e.g., chores, sports, walking) in the past month.^33^ Physical activity was then categorized into the following categories: poor (no physical activity per week), intermediate (1-149 min/week moderate intensity or 1-74 minute/week moderate/vigorous activity), and ideal (150+ min/week moderate intensity exercise or 75+ min/week vigorous intensity activity). Further, we included systolic blood pressure (mmHg; measured after 5 minutes of seated rest and by taking the average of the second and third measurements), fasting glucose levels (mg/dL), and total cholesterol levels (mg/dL) as covariates in each of our models.^33^

#### Cardiovascular Disease Risk

Based on previous research underpinning its importance,^20^ we used two outcome variables to measure cardiovascular risk in the context of this examination: carotid intima-media thickness (cIMT) and carotid plaque scores. Each measure is described in detail below.

**cIMT.** Carotid intima-media thickness (cIMT) was measured with participants in the supine position. B-mode ultrasound images of the left and right carotid artery were obtained and measured by the University of Wisconsin Atherosclerosis Imaging Research Program MESA Carotid Ultrasound Reading Center.^34^ Quality assurance measures and full measurement details are described elsewhere.^20,34^ Briefly, the far walls of the carotid arteries were measured in triplicate and cIMT reported as the mean of the mean left and mean right far wall distal carotid artery wall thickness. cIMT images were measured using the GE 700 Logic ultrasound system and a M12L MHz transducer (General Electric Medical Systems, Waukesha, WI, USA).

**Carotid plaque.** Presence of carotid plaque was indicated by abnormal wall thickness (i.e., IMT > 1.5mm) or by focal thickening of the surrounding IMT by >50%.^16,27,35^ Plaque scores were obtained from 12 carotid ultrasound images with a score ranging from 0 to 12 (i.e., if a segment met the aforementioned criteria, they were given a score of 1 and the scores were summed across the 12 segments). Full measurement details have been described previously.^16,17,20,34^ The score was calculated by assigning one point for each plaque in each segment on each side (i.e., the common carotid artery near wall, common carotid artery fall wall, carotid bulb near wall, carotid bulb far wall, internal carotid artery near all, internal carotid artery far wall). Points were assigned for both the right and left sides which gives a maximum plaque score of 12 and a minimum plaque score of 0. Each measurement was conducted at the University of Wisconsin Carotid Ultrasound Reading Center.

#### Discrimination

To assess exposure to discrimination, participants completed the Everyday Discrimination Scale (EDS).^36^ The EDS is a 9-item self-reported questionnaire on day-to-day experiences of discrimination ranging on a Likert scale from 1-6 “almost every day”, “at least once a week”, “a few times a month”, “a few times a year”, “Less than once a year”, and “never”. Participants are first asked “*In your day-to-day life, how often do any of the following things happen to you?*” and then asked a series of follow up questions pertaining to discriminatory experiences. An example item includes “*You are treated with less courtesy than other people are.*” Responses to the six items were averaged for the purposes of this study, for a total EDS score ranging from 1 (no discrimination) to 6 (heavy discrimination).

#### Current Cigarette Smoking Behavior

Our mediator of interest was current cigarette use, which we defined as one’s current cigarette smoking status. Smoking status was assessed by asking about one’s current cigarette smoking status. First, participants were asked if they had ever smoked more than 100 cigarettes in their lifetime (yes/no). Those who indicated that they had smoked at least 100 cigarettes in their lifetime were then asked if they had smoked cigarettes within the past 30 days (yes/no). We characterized smoking status as never having smoked cigarettes (0; those who reported never having smoked at least 100 cigarettes in their lifetime), formerly smoked cigarettes (1; those who reported having smoked at least 100 cigarettes in their lifetime but who did not report smoking within the past 30 days), and currently smoking cigarettes (2; those who reported smoking cigarettes within the past 30 days). In a sensitivity analysis we measured the number of cigarettes that current smokers smoke each day, on average, due to its strong association with discrimination and CVD risk. Those who reported never smoking or who reported formerly smoking were coded as smoking 0 cigarettes per day. Due to a small percentage of participants reporting over 200 cigarettes per day, we performed a 95% winsorization on the average number of cigarettes smoked per day variable prior to analysis.^37^

### Data Analysis

The two cardiovascular risk outcomes (i.e., cIMT and plaque scores) were modeled separately for the purposes of this study. We first observed demographic characteristics for both females and males for both the cIMT and carotid plaque score study samples. Then, bivariate statistics were used (e.g., chi-squared tests, t-tests) in order to examine differences in the main study variables by sex assigned at birth. We then ran eight moderated mediation models (four models were run for cigarette smoking status and four models were run for average number of cigarettes smoked per day) using the PROCESS version 4.0 macro.^38^ The PROCESS macro is a statistical tool that allows regression analyses to contain various combinations of mediators, moderators, and covariates.^9^ PROCESS macro model 59 with 5,000 bias-correct samples was used to test the moderation and mediation effects in both unconditional direct and indirect ways. Males and females were analyzed separately. In each of our models, we tested for the mediating role of cigarette use in the association between discrimination and cardiovascular risk. Further, we examined potential differences in the mediating role of cigarette use in the association between discrimination and cardiovascular risk by adding race and ethnicity as a moderator of this association within each of our models. Therefore, the direct and indirect associations between discrimination and cIMT and plaque were tested in each model. cIMT and plaque mediated by cigarette use and moderated by race/ethnicity were also investigated in the models while controlling for the possible demographic confounders. The associations were inferred significant at *p<*.05. The indirect association was inferred to be significant at the .05 level when the 95% confidence interval did not include zero. Hypothesis testing for all analyses were performed using SPSS version 27, and univariate and bivariate statistics were conducted using Stata version 17.

## Results

Sociodemographic and clinical characteristics for the cIMT and carotid plaque samples are presented in Tables 1 and 2, respectively. With respect to the cIMT sample, the study population comprised of 2,430 females (*M*_age_=61.63, SD=10.16, range=45 to 84) and 2,180 males (*M*_age_=61.71, SD=10.09, range=45 to 84). No differences were observed between females and males with respect to age, race and ethnicity, systolic blood pressure, and everyday discrimination exposure. However, there were differences in the distribution of cigarette smoking status (χ^2^(2)=181.62, *p*<.001) and physical activity levels (χ^2^(2)=30.71, *p*<.001) between females and males. Moreover, men had higher fasting glucose levels as compared to women (t=-6.96, *p*<.001) whereas females had higher total cholesterol levels as compared to males (t=11.60, *p*<.001). Finally, males had higher mean cIMT scores compared to females (t=-9.09, *p*<.001).

**Table 1.** Sociodemographic and clinical characteristics of the cIMT study sample with cigarette smoking status as the mediator.

|  |  |  |  |  |
| --- | --- | --- | --- | --- |
| | Females (n=2,430) | Males (n=2,180) | $\chi^2 / t$ | *p* |
|  | *M*(SD) / %(n) | *M*(SD) / %(n) |  |  |
| Age | 61.63(10.16) | 61.71(10.09) | -.26 | .79 |
| Race and Ethnicity |  |  | 6.54 | .09 |
| White | 37.86(920) | 38.76(845) |  |  |
| Chinese-American | 12.47(303) | 12.71(277) |  |  |
| African-American | 27.94(679) | 24.82(541) |  |  |
| Hispanic | 21.73(528) | 23.72(517) |  |  |
| Cigarette Smoking Status |  |  | 181.62 | <.001 |
| Never | 59.05(1,435) | 39.91(870) |  |  |
| Former | 28.48(692) | 45.87(1,000) |  |  |
| Current | 12.47(303) | 14.22(310) |  |  |
| Physical Activity |  |  | 30.71 | <.001 |
| Poor | 23.95(582) | 21.47(468) |  |  |
| Intermediate | 20.21(491) | 15.14(330) |  |  |
| Ideal | 55.84(1,357) | 63.39(1,382) |  |  |
| Systolic Blood Pressure (mmHg) | 126.28(22.94) | 125.85(19.32) | .69 | .49 |
| Fasting Glucose (mg/dL) | 93.56(26.03) | 99.52(32.04) | -6.96 | <.001 |
| Total Cholesterol (mg/dL) | 200.02(35.23) | 188.03(34.80) | 11.60 | <.001 |
| Everyday Discrimination | 1.60(.65) | 1.64(.68) | -1.59 | .11 |
| Mean cIMT | .75(.18) | .80(.20) | -9.09 | <.001 |

**Table 2.** Sociodemographic and clinical characteristics of the plaque study sample with cigarette smoking status as the mediator.

| | Females (n=2,755) | Males (n=2,446) | $\chi^2 / t$ | *p* |
| --- | --- | --- | --- | --- |
|  | *M*(SD) / %(n) | *M*(SD) / %(n) |  |  |
| Age | 61.74(10.10) | 61.80(10.06) | -.18 | .86 |
| Race and Ethnicity |  |  | 9.79 | .02 |
| White | 37.46(1,032) | 39.74(972) |  |  |
| Chinese-American | 11.41(328) | 12.26(300) |  |  |
| African-American | 28.82(794) | 24.98(611) |  |  |
| Hispanic | 21.81(601) | 23.02(563) |  |  |
| Cigarette Smoking Status |  |  | 192.71 | <.001 |
| Never | 59.09(1,628) | 40.27(985) |  |  |
| Former | 29.00(799) | 45.50(1,113) |  |  |
| Current | 11.91(328) | 14.23(348) |  |  |
| Physical Activity |  |  | 34.18 | <.001 |
| Poor | 24.36(671) | 21.63(529) |  |  |
| Intermediate | 20.22(557) | 15.33(375) |  |  |
| Ideal | 55.43(1,527) | 63.04(1,542) |  |  |
| Systolic Blood Pressure (mmHg) | 126.89(23.16) | 125.85(19.37) | 1.74 | .08 |
| Fasting Glucose (mg/dL) | 94.68(27.36) | 99.94(32.55) | -6.33 | <.001 |
| Total Cholesterol (mg/dL) | 199.75(35.56) | 187.78(34.67) | 12.26 | <.001 |
| Everyday Discrimination | 1.59(.64) | 1.64(.68) | -2.57 | .01 |
| Mean Plaque | 1.17(1.68) | 1.51(1.99) | -6.69 | <.001 |

In terms of the carotid plaque sample, the study population comprised of 2,755 females (*M*_age_=61.74, SD=10.10, range=44 to 84) and 2,446 males (*M*_age_=61.80, SD=10.06, range=44 to 84). As shown in Table 2, no differences were observed between females and males with respect to age and systolic blood pressure. However, there were differences in the distribution of race and ethnicity (χ^2^(2)=9.79, *p*=.02), cigarette smoking status (χ^2^(2)=192.71, *p*<.001), and in physical activity levels (χ^2^(2)=34.18, *p*<.001) between females and males. In terms of fasting glucose, men had higher fasting glucose levels as compared to women (t=-6.33, *p*<.001). On the other hand, females had higher total cholesterol levels as compared to males (t=12.26, *p*<.001). With respect to everyday discrimination, males reported higher everyday discrimination as compared to females (t=-2.57, *p*=.01). Lastly, males had higher mean plaque scores as compared to females (t=-6.69, *p*<.001).

### Females

**cIMT.** The omnibus test of the model predicting cIMT using cigarette smoking status as the mediating variable among female participants was significant, (*F*(17, 2412)=68.12, *p*<.001) and explained 56.96% of the variance in mean cIMT among female participants. However, our study hypotheses were not supported among females in terms of cIMT. First, we did not find enough evidence to suggest that cigarette use mediated the association between everyday discrimination and cIMT among females. This was evidenced by the index of moderated mediation not reaching significance for Chinese (index = −.002, CI = −.007, .001), Black (index = −.001, CI = −.003, .001), and Hispanic (index = .001, CI = −.002, .004) females in comparison to White females. Further, when examining the conditional indirect association of everyday discrimination on cIMT among the racial and ethnic groups, they did not reach statistical significance among our sample of female participants.

**Carotid Plaque.** The omnibus test of the model predicting carotid plaque scores among females was significant, (*F*(17, 2737)=38.11, *p*<.001) and explained 43.75% of the variance in carotid plaque score among female participants. Figure 2 displays the direct and mediated moderated associations between the study variables among female participants for mean plaque scores. While we controlled for total age, cholesterol, systolic blood pressure, fasting glucose, and physical activity, these variables are not included in the figure. Our study hypotheses were not fully supported among females as we did not find enough evidence to suggest that cigarette use mediated the association between everyday discrimination and carotid plaque score as the index of moderated mediation did not reach significance for Chinese (index = −.03, CI = −.08, .01), Black (index = −.01, CI = −.06, .03), or Hispanic (index = .01, CI = −.04, .07) females in comparison to White females. However, we did find a significant direct association of cigarette smoking among female participants such that those who smoked previously or currently had higher carotid plaque scores than those who never smoked (*b*=.45, *p*<.001). Further, we found a significant conditional indirect association of everyday discrimination on carotid plaque score among Hispanic females (see Table 3). Thus, for Hispanic females, the association between discrimination and carotid plaque was mediated through cigarette smoking in such a way that, compared to Hispanic females who experienced less discrimination and thus more likely to be in the non-smoking group, more discrimination was associated with more cigarette use, and, in turn, higher mean plaque scores.

**Figure 2.**
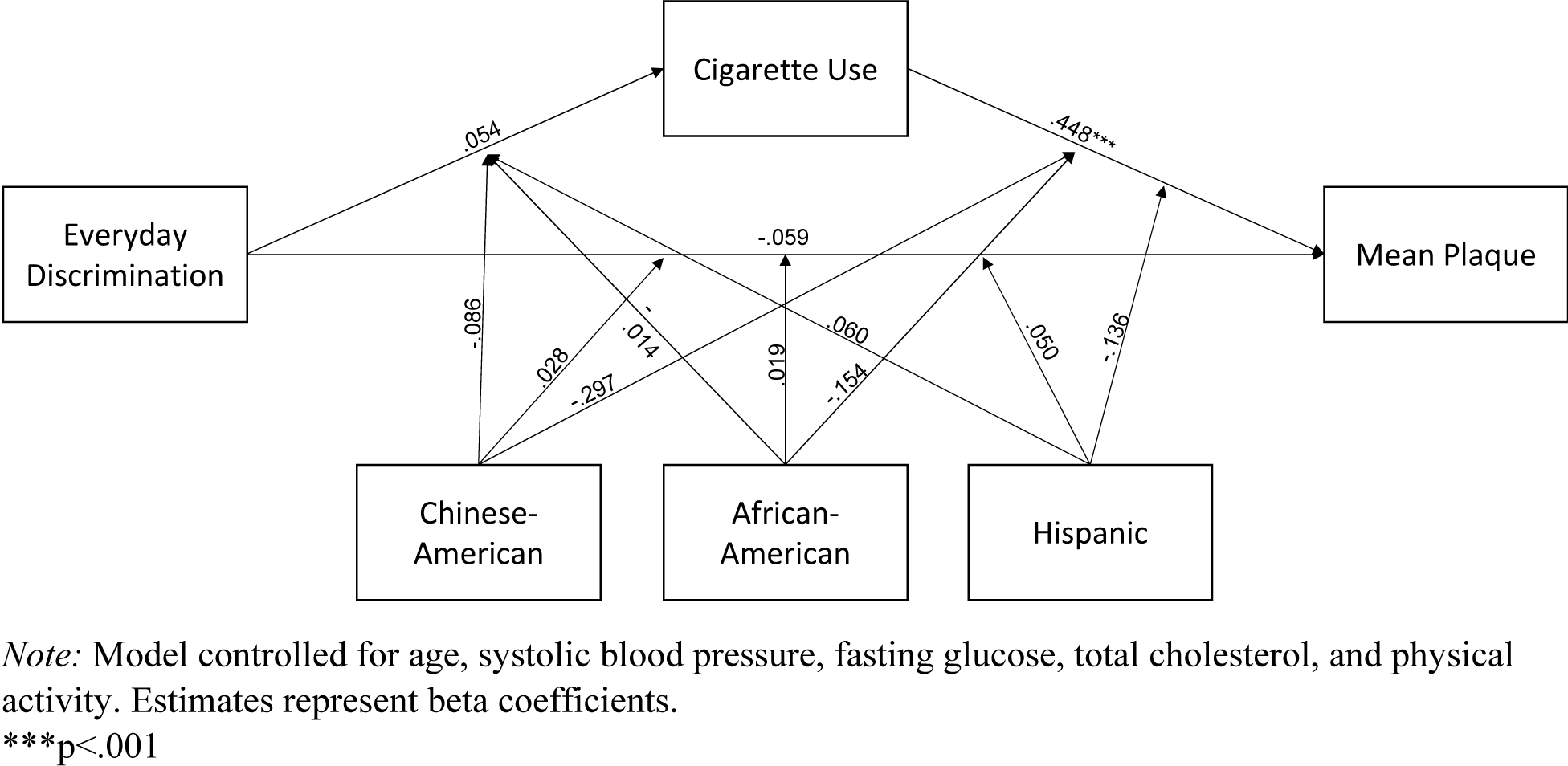
Mediated moderation model predicting mean plaque via current cigarette smoking status moderated by race and ethnicity among females

**Table 3.** Conditional indirect effects of everyday discrimination on mean plaque score among females with cigarette smoking status as the mediator.

|  | Effect | SE | CI [LLCI, ULCI] |
| --- | --- | --- | --- |
| White | .024 | .020 | -.01, .06 |
| Chinese-American | -.005 | .016 | -.048, .015 |
| African-American | .012 | .012 | -.009, .038 |
| Hispanic | .036* | .019 | .007, .080 |

\*p<.05

### Males

**cIMT.** The omnibus test of the model predicting mean cIMT with current smoking status as the mediating variable among male participants was significant, (*F*(17, 2162)=43.13, *p*<.001), and explained 50.33% of the variation in cIMT among male participants. Figure 3 displays the direct and mediated moderated associations between the study variables among male participants for mean cIMT scores. Our hypothesis that cigarette use would mediate the association between everyday discrimination and mean cIMT among male participants was not fully supported as the index of moderated mediation did not reach significance among Chinese (index = −.01, CI = −.02, .002), Black (index = −.0003, CI = −.003, .002), or Hispanic (index = .002, CI = −.002, .006) men in comparison to White men. We found evidence to suggest that everyday discrimination was associated with cigarette use among our sample of male participants such that experiencing more discrimination was associated with a greater likelihood of smoking cigarettes as compared to those who experienced less discrimination (*b*=.09, *p*=.03). With respect to mean cIMT, we did not find evidence to suggest a significant direct association between everyday discrimination and mean cIMT among males. However, we found evidence for a significant conditional indirect association of everyday discrimination on mean cIMT among Hispanic males (see Table 4). Thus, for Hispanic males, the association between discrimination and mean plaque was mediated through cigarette smoking in such a way that greater exposure to discrimination was associated with higher levels of smoking which, in turn, was associated with higher mean cIMT. Significant conditional associations of cigarette use were observed among Chinese-American and Hispanic males such that higher levels of cigarette use was associated with an increase in mean cIMT. The slope for cigarette use on mean cIMT was steepest among Chinese-American males (*b*=.05, *p*<.01) followed by Hispanic males (*b*=.03, *p*=.01).

**Figure 3.**
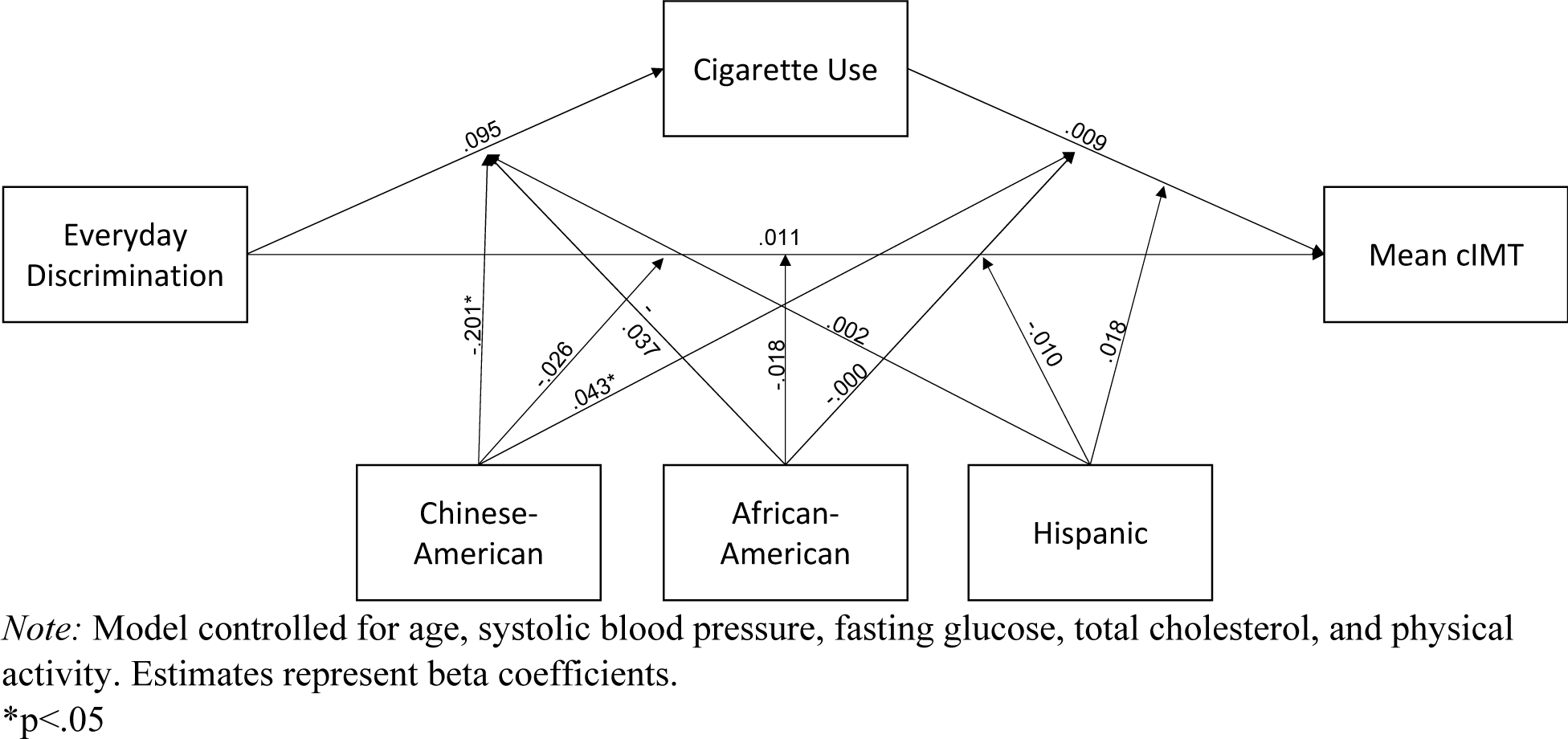
Mediated moderation model predicting mean cIMT via current cigarette smoking status moderated by race and ethnicity among males

**Table 4.** Conditional indirect effects of everyday discrimination on mean cIMT score among males with cigarette smoking status as the mediator.

|  | Effect | SE | CI [LLCI, ULCI] |
| --- | --- | --- | --- |
| White | .001 | .001 | -.001, .003 |
| Chinese-American | -.006 | .004 | -.015, .002 |
| African-American | .001 | .001 | -.001, .003 |
| Hispanic | .003* | .002 | .000, .007 |

\* $p < .05$

**Carotid plaque.** The omnibus test of the model predicting carotid plaque score among male participants was significant, (*F*(17, 2428)=32.62, *p*<.001), and explained 43.12% of the variation in plaque among male participants. Our hypothesis that cigarette use would mediate the association between everyday discrimination and mean plaque among males was not fully supported. While the index of moderated mediation did not reach significance among Black (index = −.02, CI = −.07, .03) or Hispanic (index = −.007, CI = −.06, .04) men in comparison to White men, the index of moderated mediation was significant among Chinese men (index = −.04, CI = −.09, −.001). Figure 4 displays the direct and mediated moderated associations between the study variables among male participants for carotid plaque scores. We found that everyday discrimination was associated with cigarette use among males (*b*=.09, *p*=.02) whereas everyday discrimination was not associated with mean plaque among males. However, we found significant conditional indirect associations of everyday discrimination on carotid plaque among White and Hispanic males (see Table 5). Among both White and Hispanic males, greater levels of everyday discrimination, as compared to experiencing less everyday discrimination, was associated with a greater likelihood of smoking, which, in turn, was associated with higher carotid plaque scores.

**Figure 4.**
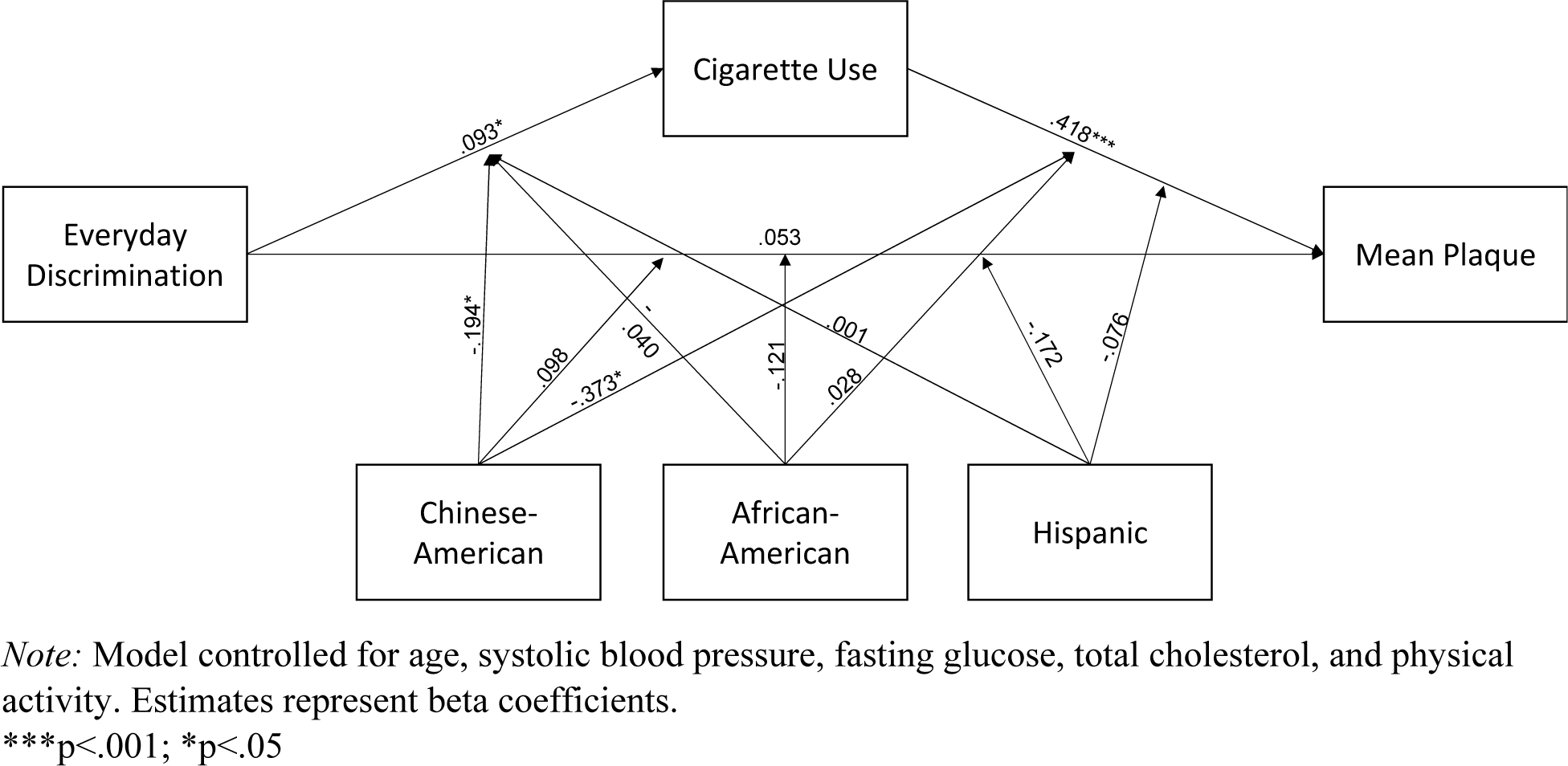
Mediated moderation model predicting mean plaque via current cigarette smoking status moderated by race and ethnicity among males

**Table 5.**
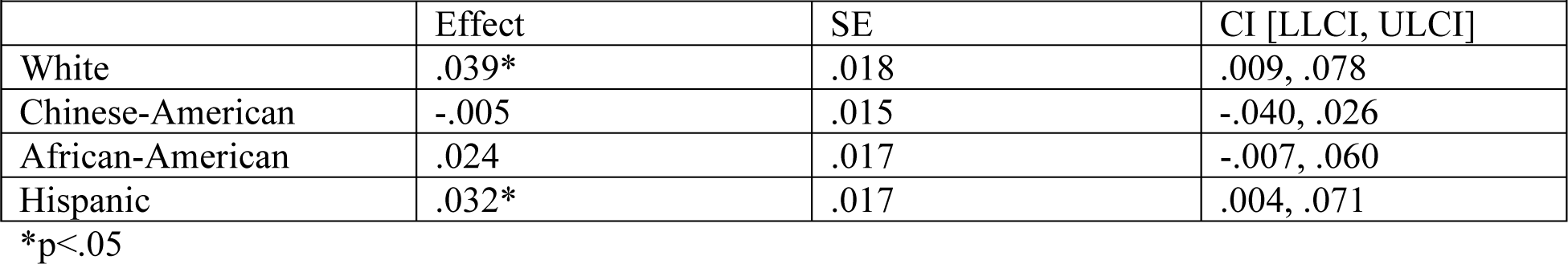
Conditional indirect effects of everyday discrimination on mean plaque score among males with cigarette smoking status as the mediator.

### Sensitivity Analyses

Sociodemographic and clinical characteristics of the analytic samples used in sensitivity analyses are presented in Supplemental Tables 1 and 2 for cIMT and carotid plaque, respectively.

**cIMT.** We did not find enough evidence to suggest that cigarette use mediated the association between everyday discrimination and cIMT among females. This was evidenced by the index of moderated mediation not reaching significance for Chinese (index = −.002, CI = −.017, .001), Black (index = −.000, CI = −.002, .001), and Hispanic (index = −.000, CI = −.002, .002) females in comparison to white females. Further, when examining the conditional indirect associations of everyday discrimination on cIMT among the racial and ethnic groups, they did not reach statistical significance among our sample of female participants.

Among males, our hypothesis that cigarette use would mediate the association between everyday discrimination and mean cIMT among male participants was not fully supported as the index of moderated mediation did not reach significance among Chinese (index = −.006, CI = −.016, .000), Black (index = .000, CI = −.002, .002), or Hispanic (index = .000, CI = −.002, .002) men in comparison to white men. We found evidence to suggest that everyday discrimination was associated with cigarette use among our sample of male participants such that experiencing more discrimination was associated with a smoking a greater number of cigarettes per day as compared to those who experienced less discrimination (*b*=.70, *p*=.01). Further, as compared to white men, Chinese (*b*=2.08, *p*=.017) and Black (*b*=1.58, *p*=.011) men smoked more cigarettes per day, on average. With respect to mean cIMT, we did not find evidence to suggest a significant direct association between everyday discrimination and mean cIMT among males. However, we found evidence for a significant conditional indirect association of everyday discrimination on mean cIMT among Chinese males (see Supplemental Table 3).

**Carotid plaque.** Among females, we did not find enough evidence to suggest that cigarette use mediated the association between everyday discrimination and carotid plaque score as the index of moderated mediation did not reach significance for Chinese (index = −.014, CI = −.067, .030), Black (index = −.010, CI = −.050, .028), or Hispanic (index = −.003, CI = −.047, .047) females in comparison to white females. However, we did find a significant direct association of cigarette smoking among female participants such that those who smoked previously or currently had higher carotid plaque scores than those who never smoked (*b*=.07, *p*<.001).

Among men, the index of moderated mediation did not reach significance among Chinese (index=−.006, CI=−.053, .040), Black (index = −.012, CI = −.057, .031) or Hispanic (index = −.022, CI = −.062, .016) men in comparison to white men. We found that everyday discrimination was associated with cigarette use among males (*b*=.61, *p*=.01) whereas everyday discrimination was not associated with mean plaque among males. Further, Chinese (*b*=2.06, *p*=.01) and Black (*b*=1.18, *p*=.04) men smoked more cigarettes per day than white men, on average. Supplemental Figure 2 displays the direct and mediated moderated associations between the study variables among male participants for carotid plaque scores. We also found that men who smoke more cigarettes per day have higher carotid plaque scores on average (*b*=.04, *p*<.01). Lastly, we also observed a significant indirect association of everyday discrimination on carotid plaque scores for white men such that those who experienced more discrimination had higher plaque scores on average than those who experienced less discrimination (see Supplemental Table 4).

## Discussion

In this study, we sought to examine the mediating association of current cigarette smoking status on the association between everyday discrimination and mean cIMT and carotid plaque burden among female and male individuals residing in the United States utilizing data from the Multi-Ethnic Study of Atherosclerosis (MESA). Further, we sought to explore differences in these associations by race and ethnicity. We hypothesized that cigarette smoking status would mediate the association between everyday discrimination and CVD risk among both males and females, and that these associations would differ according to racial and ethnic identity. Specifically, we hypothesized that there would be a more pronounced association between everyday discrimination, cigarette smoking status, and CVD risk among racial and ethnic minorities as compared to whites (i.e., more everyday discrimination among racial and ethnic minorities would be associated with greater levels of smoking which, in turn, would be associated with greater CVD risk). However, overall, our hypotheses were not supported.

For females, we did not find evidence to suggest that cigarette smoking status mediated the association between exposure to everyday discrimination and mean cIMT. Further, the conditional indirect effects of everyday discrimination on mean cIMT across the racial and ethnic groups did not reach significance among females. Similarly, we did not find enough evidence to suggest that cigarette smoking status mediated the association between everyday discrimination and carotid plaque score among females. However, we did observe a significant conditional indirect association of everyday discrimination on carotid plaque score, specifically among Hispanic females such that compared to those who experienced less everyday discrimination, those who experienced more were more likely to use cigarettes which, in turn, was associated with higher mean plaque scores among Hispanic females. Overall, our finding that Hispanic women may experience greater CVD risk via discrimination is supported in previous research.^39^ For instance, Brondolo, Libby, Denton, Thompson, Beatty, Schwartz, Sweeney, Tobin, Cassells, Pickering and Gerin^40^ found that perceived racism was associated with increased nocturnal ambulatory blood pressure (ABP) among a sample of Blacks and Hispanics. However, in terms of racial and ethnic differences among women in the association between discrimination and cardiovascular risk, evidence is mixed. Indeed, using data from 2,180 women who participated in the Study of Women’s Health Across the Nation (SWAN), Moody, Chang, Pantesco, Darden, Lewis, Brown, Bromberger and Matthews^41^ found that although there was a positive association between discrimination and blood pressure there were no observed racial or ethnic differences in this association across those who identified as White, Black, Chinese, Japanese, or Hispanic. However, the association between discrimination and CVD risk may vary greatly depending on how CVD risk is measured.^42^ For instance, in a systematic review examining discrimination and cardiovascular risk, Panza, Puhl, Taylor, Zaleski, Livingston and Pescatello^42^ found that, of 43 studies examining blood pressure as an outcome in relation to racial discrimination, 11 of them did not observe an association between discrimination and blood pressure. However, when examining CVD risk outcomes such as heart rate variability (HRV), each of the 8 studies found a positive association between racial discrimination and HRV. Thus, more work needs to be done with more robust measures of CVD risk to better understand the complex association between discrimination and CVD risk across diverse samples of women.

We found some support for our hypotheses among males. Among Hispanic males, we found that greater levels of everyday discrimination were associated with higher levels of smoking which, in turn, was associated with greater mean cIMT. Further, we found evidence for significant conditional associations of cigarette use among Chinese-American and Hispanic males suggesting that more cigarette use was associated with increased mean cIMT. In terms of carotid plaque scores among males, we found significant conditional indirect associations of everyday discrimination on carotid plaque scores among White and Hispanic males such that greater exposure to everyday discrimination was associated with smoking which, in turn, was associated with higher mean plaque scores among these racial/ethnic groups. While our hypotheses regarding the mediating role of cigarette use on the association between everyday discrimination and cardiovascular risk indices were not fully supported, we explore potential explanations for our findings and suggest future directions in this area of inquiry below.

Among males, we found evidence to suggest that experiencing more everyday discrimination was associated with greater cigarette use which, in turn, was associated with greater mean cIMT among Hispanics. Further, we found differences in the conditional effect of cigarette use on mean cIMT by race and ethnicity among males. In particular, we observed the strongest association between cigarette use and mean CIMT among Chinese-American males, followed by Hispanic males. In terms of carotid plaque, we found that greater exposure to everyday discrimination was associated with a greater likelihood of smoking, which, in turn, was associated with higher levels of mean plaque specifically among White and Hispanic males. Previous research may help to partially explain our study findings. In general, research demonstrates differences in cardiovascular risk among men by race and ethnicity. For instance, African-American and Asian American men are more likely to exhibit signs of cardiovascular risk such as heightened blood pressure as compared to White men.^43,44^ However, determining difference specifically based on the intersection or race and gender more complex. Bey, Jesdale, Forrester, Person and Kiefe^45^ sought to examine differences in the association between intersectional discrimination (i.e., racial and/or gender discrimination) and cardiovascular health among 5,114 White and Black individuals who participated in the Coronary Artery Risk Development in Young Adults (CARDIA) study. These authors found that the association between exposure to discrimination and cardiovascular health (i.e., Life’s Simple 7 Cardiovascular Health score) differed between White and Black men. In particular, the authors found no association between exposure to either racial or gender discrimination, or the combination of both, and cardiovascular health among Black men. Among White men, however, experiencing racial discrimination was associated with an increase in the cardiovascular health score whereas the experience of both racial and gender discrimination was associated with a decrease in the cardiovascular health score. Thus, there may be key differences in the appraisal of discrimination among men across racial and ethnic groups depending on severity and type of discrimination experienced that warrants further exploration.

We propose that there are several explanations for why we did not observe an association between discrimination and cardiovascular risk across varying racial and ethnic groups of women (e.g., African American, Chinese American) that includes understanding differences in the cultural experiences of women of different races and ethnicities that may differentially influence that ways in which they internalize, perceive, and cope with discrimination events. Having a better understanding of these nuanced mechanisms could explain the ways in which discrimination is associated with cardiovascular risk. For instance, Johnson, McCloyn and Sims^46^ examined the role of high-effort coping style (i.e., a coping mechanism that utilizes significant cognitive and emotional resources) in the association between everyday discrimination and cardiovascular risk as measured by allostatic load among African-Americans. These authors found evidence to suggest that the association between everyday discrimination and allostatic load differed by coping style such that those who experienced everyday discrimination and who engaged in high-effort coping as a response were more likely to have higher allostatic load than those who did not engage in high-effort coping. Thus, there may be key cultural differences across women of varying races and ethnicities that attenuates or amplifies the impact of discrimination on CVD risk which should be addressed in future research.

Next, it may be that racial and ethnic minority women and men become habituated to the effects of experiences of perceived discrimination over time. In other words, individuals may develop positive coping mechanisms (e.g., social support) to cope with experiences of discrimination which may, in turn, habituate the negative effects of discrimination on physical health over time. Some research exists in support of this hypothesis; however, this needs to be explored with further research. Indeed, Kondrat, Sullivan, Wilkins, Barrett and Beerbower^47^ found evidence supporting a mediation model suggesting that exposure to discrimination led to increased social support behaviors which, in turn, was associated with better overall mental health. To better understand potential explanatory mechanisms of the relationship between discrimination, social support, and cardiovascular risk among racial and ethnic minorities, researchers may wish to utilize longitudinal study designs. For instance, much of the extant research examining social determinants of health and CVD risk focus on direct pathways between the two. However, there may be key mediators at play within these associations. More research needs to be conducted on how individuals appraise perceived discrimination events and how, in turn, individuals cope with these appraisals of discrimination. For instance, if an individual experiences discrimination, do they experience increased psychological stress, which, in turn, impacts social support behaviors and/or other coping mechanisms such as substance use? Complex longitudinal study designs could aid in our understanding of how the potential mediating role of coping mechanisms may explain why we did not observe differences in the association between everyday discrimination, cigarette use, and mean cIMT and mean plaque across different racial/ethnic identities (e.g., African Americans).

Although our hypotheses were not fully supported among our study sample, our findings highlight several implications and areas for future inquiry. First, our findings pinpoint a need for an intersectional framing of research analyses when examining the association between social determinants of health such as discrimination and cardiovascular risk among varying racial and ethnic groups. As observed in our results, we found differential impacts, or lack thereof, of everyday discrimination and cigarette smoking on cardiovascular risk across females and males. While our findings did not fully align with our conceptual model adapted from Lewis, Lampert, Charles and Katz^22^, there are key mediators along the causal pathway discussed in this model that need to be expanded upon in future research in an intersectional manner. So, for instance, there may be key cultural differences in the experiences and appraisals of experiences of discrimination and how, these in turn, impact coping behaviors and patterns of physical health across racial and ethnic groups which we were not able to examine in the present study. These differences may be observed across and within social identities such as race and ethnicity and gender identity due to differences in factors such as culture, familial/social relationships, and social norms. For instance, one study examining the impact of discrimination on mental health among Asian American and Latinx college students found that, while both groups experienced similar levels of discrimination, the type of discrimination and reactions to discriminatory experiences differed.^48^ Compared to Asian American students, Latinx students were more likely to be accused of wrongdoing such as cheating on an exam or breaking the law and were more likely to appraise their experiences with discrimination as stressful. While we were not able to capture such information in this secondary data analysis, we are one of the first to examine how social determinants of health influence health behaviors and cardiovascular risk (namely, plaque burden) among a racially/ethnically diverse study sample in the United States. Our results suggest a complicated understanding of the ways in which social determinants of health influence CVD risk across different social groups (i.e., biological sex and race/ethnicity). However, there remains a critical need to understand *how* discrimination is experienced, in what ways these experiences are appraised, and how, in turn, individuals cope across varying social identities and how these all intertwine to influence patterns of physical health.

With respect to the sensitivity analyses, there were a few notable differences. With respect to cIMT, when including average number of cigarettes per day as the mediating variable we observed a significant conditional indirect effect of discrimination on cIMT among Chinese males such that greater levels of discrimination was associated with lower cIMT. Further, in our carotid plaque models, we no longer observed the conditional indirect effect of discrimination on plaque among both Hispanic women and men. Overall, however, sensitivity analyses suggest that heightened cigarette use was associated with higher plaque scores among both women and men and that experiencing more discrimination was associated with greater cigarette use overall among men. While our results regarding racial and ethnic differences in the association between discrimination, cigarette use, and cardiovascular risk slightly differ when examining number of cigarettes smoked per day, we may have been limited in that the majority of the sample reported smoking 0 cigarettes per day.

This study is not without limitations. First, our study was cross-sectional in nature as we used data from the baseline examination of MESA. This limits our ability to fully examine the causal pathways posited in our mediation model (e.g., it is unknown if smoking behaviors came after exposure to everyday discrimination and so on). Nevertheless, this is one of the first studies to examine the mediating impact of cigarette consumption in the association between everyday discrimination and cardiovascular risk among a racially and ethnically diverse group of women and men in the United States. While MESA collects cardiometabolic health data across multiple years, we were limited due to the measurement of our outcomes (i.e., cIMT and carotid plaque) being limited in future waves which would have affected our power to detect a significant pattern. However, our cross-sectional approach may be justified within the context of this examination as our research is grounded by a strong causal theoretical framework that describes the way in which discrimination influences psychological processes that then influences coping behaviors such as smoking which, in turn, may influence cardiovascular risk over time.^22^ Further, the interpretation of the discrimination variable relates to experiences of discrimination on a day-to-day basis and implies that discrimination occurred before smoking behaviors (which we measured as current cigarette smoking behavior). In such cases, cross-sectional mediation analyses can be appropriate as there is both a strong causal framework underpinning our study and the interpretation of our discrimination and smoking variables implies that one occurs before the other.^49^ Second, the MESA cohort is not a representative sample of individuals who reside in the United States; thus, our findings may not be generalizable to the United States as a whole. There may be key differences to consider by geographic location in the association between discrimination and cardiovascular risk that we were unable to explore in this examination. Third, cIMT and carotid plaque are just two measures of cardiovascular risk. There may be other measures, or combinations of measures (e.g., IL-6, allostatic load), that may more fully capture cardiovascular risk. Nevertheless, research demonstrates that cIMT and carotid plaque scores are reliable indicators of cardiovascular risk.^15,50,51^ Lastly, MESA excluded individuals with known cardiovascular disease baseline. Thus, our sample may be limited in our examination of smoking and cardiovascular health.

## Data Availability

Data for the Multi-Ethnic Study of Atherosclerosis are available upon approved application at: https://www.mesa-nhlbi.org/.

## Future Directions

There are several areas to highlight for future research that are inspired by the findings of this study. First, there needs to be more work specifically focused on understanding the varying cultural experiences that may shape experiences of discrimination and resultant CVD risk. Our findings highlight the need to examine these associations among traditionally understudied racial and ethnic minority populations including Chinese American individuals. Nevertheless, it remains unclear what the important differences are in experiences of discrimination at different axes of gender and race.^52^ Thus, there is a key need for more mechanistic research that more wholly explains the association between discrimination and cardiovascular risk while examining these important intersections of racial and gender identity. For instance, examining patterns of skin-deep resilience and coping mechanisms such as social support or denial of one’s discrimination experiences are important areas to consider in future work and should be explored using an intersectional lens. Further, current research suggests that social determinants of health, including exposure to racial discrimination, may explain racial and ethnic differences observed in cardiovascular risk ^3^. While some recent work demonstrates that social conditions such as discrimination may not infer stress that influences physiological processes such as cardiovascular risk, the evidence is mixed and needs further expansion with future work including longitudinal study designs.^53^ Indeed, more work is needed regarding how different social groups experience discrimination and how the appraisals of these experiences influence health behaviors and physical health across the life-course. Overall, utilizing a mechanistic approach and discovering specific differences in social determinants of health based on key intersecting identities and experiences will add in the creation of more precise cardiovascular risk prevention interventions.

